# The BRoccoli In Osteoarthritis (BRIO study) - A randomised controlled feasibility trial to examine the potential protective effect of broccoli bioactives, (specifically sulforaphane), on osteoarthritis

**DOI:** 10.1101/2024.06.20.24309233

**Authors:** Rose K Davidson, Laura Watts, Gemma Beasy, Shikha Saha, Paul Kroon, Aedin Cassidy, Allan Clark, William Fraser, Iain Mcnamara, Sarah R Kingsbury, Philip G Conaghan, Ian M Clark, Alex Macgregor

## Abstract

**Objective:** The Broccoli in Osteoarthritis (BRIO Study) was conducted to determine whether dietary sulforaphane (SFN), consumed as broccoli, improves pain and/or physical function in participants with knee osteoarthritis (OA). This was a proof of principle study to test the feasibility of the trial to optimise the design of an appropriately powered study.

**Design:** Two-centre, double-blind, two-arm parallel, randomised placebo-controlled, dietary intervention feasibility trial. Patients with radiographic knee osteoarthritis (Kellgren-Lawrence score 2-3), with pain of at least 4 on a scale of 0-10 were recruited. The intervention was a high glucoraphanin broccoli, (source of SFN), or a matched placebo (no SFN) soup. Pain and measures of physical function were measured at baseline, 6 and 12 weeks.

**Results:** The mean WOMAC pain score (scale 0 - 20) was decreased by 4.2 (95% CI: 1.03,7.38) following intervention, Similar patterns of improvement were observed for other pain and function outcome measures. Study data, sample collections and intervention adherence were 100% compliant except where COVID restrictions applied. Acceptability for randomisation was 100% and acceptability for the intervention was 92%. There were three related adverse events, two of which were expected.

**Conclusions:** High glucosinolate broccoli soup is a novel approach to managing OA that is widely accessible and can be used on a large scale. This study shows that it is an acceptable way of delivering dietary bioactives and has potential for therapeutic benefit. The primary outcome of pain improved in the intervention group compared to the placebo and the confidence interval encompassed the minimal clinically important difference. The data provide justification for proceeding to a large scale, appropriately powered intervention trial.

## Introduction

Treatment options for osteoarthritis (OA) are limited. Aside from surgery, there are no current safe long-term effective analgesic treatments for OA. Sensitisation of nociceptors by pro-inflammatory cytokines in the joint tissues is a suggested mechanism for OA pain, and some short-term pain relief can be gained using NSAIDs and corticosteroids, which may in part, reduce synovitis. However, the well-documented contra-indications for their long-term use are undesirable. New options are required to reduce pain and improve quality of life for people with OA.

Dietary interventions are perceived as safe and are supported by OA patients. In a study of over 1200 patients 68% of complementary and alternative medicine users explored dietary supplements to manage their disease and symptoms^1^. OA patients tend to believe diet impacts their OA symptoms and they are interested in nutritional education for self-management of their disease^2,3^. Importantly, dietary intervention has the potential to be used on a large scale.

Glucoraphanin is a cytoprotective dietary phytochemical found in the Brassicaceae family. Sulforaphane (SFN) is an isothiocyanate metabolite of glucoraphanin that can be detected in plasma following consumption of dietary sources and the action of myrosinase either from the plant itself or following breakdown/metabolism by the gut microbiota. There is a growing body of evidence that supports SFN as a new potential OA therapeutic. SFN has been shown to modulate surrogate markers of OA in vitro, ex vivo and in vivo laboratory models of OA with effects on NF-κB and Nrf2 signalling. Human feeding studies have shown that the consumption of broccoli can lead to detectable levels of SFN or its metabolites in plasma, urine, and synovial fluid, while in vitro studies show SFN can accumulate intracellularly via conjugation to glutathione giving millimolar intracellular concentrations. Additionally, SFN treatment in rodent models of arthritis have demonstrated a protective role in reducing inflammation and cartilage destruction^4–9^.

Evaluating SFN in a randomised trial requires better evidence of feasibility in humans. This study is designed as a feasibility study to determine if SFN, naturally present in broccoli, improves pain in people with knee OA.

## Methods

### Objectives

The primary objective was to determine whether dietary SFN, gained from the consumption of broccoli, improves pain in people with knee OA. Secondary objectives were to determine whether food containing SFN improves physical function in people with knee OA, estimate heterogeneity in response to the intervention, feasibility of recruitment and retention, test compliance, the sensitivity of the outcome measures, examine potential adverse effects and acceptability of randomisation, and finally to determine the sample size estimates for a subsequent longer-term trial.

### Trial design

UK-based two-centre, double-blind, two-arm parallel, random placebo-controlled feasibility trial. Trial centres were University of East Anglia/Norfolk & Norwich University Hospital (NNUH) and University of Leeds/Chapel Allerton Hospital (CAH).

### Participants

Participants had radiographic knee OA (Kellgren-Lawrence score 2-3) with moderate to severe knee pain of at least 4 on 0-10 numeric rating scale. Participants were over 50 years of age and recruited from the regions of Norfolk and Leeds in the United Kingdom. Full inclusion and exclusion criteria are in **Supplementary File 1.** Data were collected in a clinical examination setting.

### Intervention

Intervention was a 300 mL portion of high glucoraphanin (120 μmol) soup (supplied by Professor Richard Mithen, Institute of Food Research, Norwich, UK), containing base vegetables and broccoli (equivalent to three 75 g portions of broccoli a day). The placebo control was a sensory matched 300 mL portion of soup containing base vegetables only. The soups were consumed once daily on any four days per week, for 12 weeks to include one for lunch the day before a follow-up visit day. Participants underwent a washout period where a restricted diet, (devoid of isothiocyanate sources), was followed for 3 days prior to baseline visit and throughout the study thereafter. Participants were supplied with a list of restricted foods that contained isothiocyanates to avoid throughout the study (**Supplementary File 2**). Participants were allowed to use their usual medications and usage was recorded. Standard meals were supplied for consumption on the evening before a visit day.

### Randomisation

Participants were randomised 1:1 to one of two trial arms using the web-based REDCap (Research Electronic Data Capture v13.1.27) tools hosted by the Norwich Clinical Trials Unit. Allocation was stratified by centre (NNUH/CAH), sex (male/female) and age (<=60 / >60) using permuted block randomisation with randomly varying block sizes of 2 and 4. Allocation was concealed prior to randomisation to prevent treatment bias. As per SAGER guidelines male or female classification is based on biological distinction to the extent that this is possible to confirm. Sex of participants was self-reported.

### Blinding

All participants, investigators, clinical staff, and outcome assessors were blinded to the intervention throughout the study.

### Outcome Measures

The primary outcome measure was a change in the Western Ontario and McMaster (WOMAC ®) Index (WOMAC Osteoarthritis Index LK3.1) pain subscale between baseline and 12 weeks. This WOMAC was 24 self-reported questions comprising a 5-point Likert scale ((0-4) none, mild, moderate, severe, extreme). It is reported as three separate subscales for pain (scale out of 20), stiffness (scale out of 8) and function (scale out of 68). The secondary outcome measures were changes (at 6 and 12 weeks) in WOMAC pain (at 6 weeks), WOMAC physical function, WOMAC stiffness, changes in an 11-point NRS (Numerical Rating Scale)^10^ for pain and function measures, A Measure of Intermittent and Constant Osteoarthritis Pain: KNEE version 3.1 (ICOAP) and the use of rescue analgesics/NSAIDs. Plasma and urine samples for objective measures of compliance (metabolites of glucoraphanin), and validated dietary data (Food Frequency Questionnaire, and 24-hour recall (INTAKE24^11^)) were collected (data available on request).

### Sample size

This was a feasibility study where the main statistical aim is to use the data generated to inform the sample size calculation of a full trial which is powered to detect a difference between the treatments. In this setting sample size calculations based on the ability to detect a difference between the treatment arms are generally not considered appropriate. Sim and Lewis^12^ and Lancaster et al^13^ have recommended sample size of n=30 and 50 are sufficient to deliver reliable findings in this setting. Our original aim was to achieve a target sample size of 64.

The study comprised four visit days to include a screening visit, baseline visit and two follow-up visits at 6 and 12 weeks. Trial registration numbers: ISRCTN 11629849, CPMS 40910, ClinicalTrials.gov NCT03878368

### Ethics

The procedures followed were in accordance with the ethical standards of NHS Health Research Authority and Health and Care Research Wales (HCRW) and with the Helsinki Declaration of 1975, as revised in 2000. Ethical approval for the study was granted by East of England - Cambridge East Research Ethics Committee (ref 19/EE/0007), IRAS: 250371. All patients gave their full informed and written consent to participate in the study. All patient data was anonymised.

### Analysis

Descriptive statistics of outcomes are reported by randomised groups. Percentages are of non-missing values. No formal hypothesis tests will be used; however, 95% and 80% confidence intervals will be given for the estimated effect size.

For continuous questionnaire outcome measures the mean difference will be estimated and confidence interval based on a two-sample t-test and an adjusted for baseline analysis using linear regression. The number of joints will be compared using a difference of medians estimated using a generalized Hodges-Lehmann estimator.

No adjustment for the stratification factors used in the randomisation will be undertaken due to the sample size. Analysis was completed using Stata 18.0.

### Analyses of Sulforaphane and Sulforaphane conjugates

Metabolites of glucoraphanin including sulforaphane (SFN), sulforaphane-cysteine (SFN-Cys), sulforaphane-N-acetylcysteine (SFN-NAC) and erucin-NAC (E-NAC) were quantified in plasma and urine samples as previously described^14^, except where d8-sulforaphane was used an as internal standard and an Acquity UPLC, HSS T3 1.8 um, 2.1 × 100 mm column was used instead of Phenomenex® Luna 3u C18 (2) 100A 2.1 × 100 mm column for improved sensitivity.

## Results

### Study subjects

The start of recruitment to this study coincided with the COVID19 pandemic when national recruitment to non-COVID19 related trials at the two recruiting centres was periodically halted. A total of 150 patients enquired about the trial of which 65 were potentially eligible. Funding and longevity of the intervention products limited our recruitment to inviting n=37 participants to full screening appointment and consented to take part. The trial steering committee and progress group deemed this number sufficient to be likely to be sufficiently informative to address the study’s principal aims as a feasibility study. A total of 24 met the screening criteria, with randomisation delivering 17 to the control group and 7 to the intervention group. There were two participant withdrawals that consented to continuation with an intention to treat (ITT) at week 6 and one drop out at week 12, resulting in a final study participant number of n=23 for analysis.

The participant characteristics of the intervention and control groups are shown in **Table 1**.

**Table 1.**
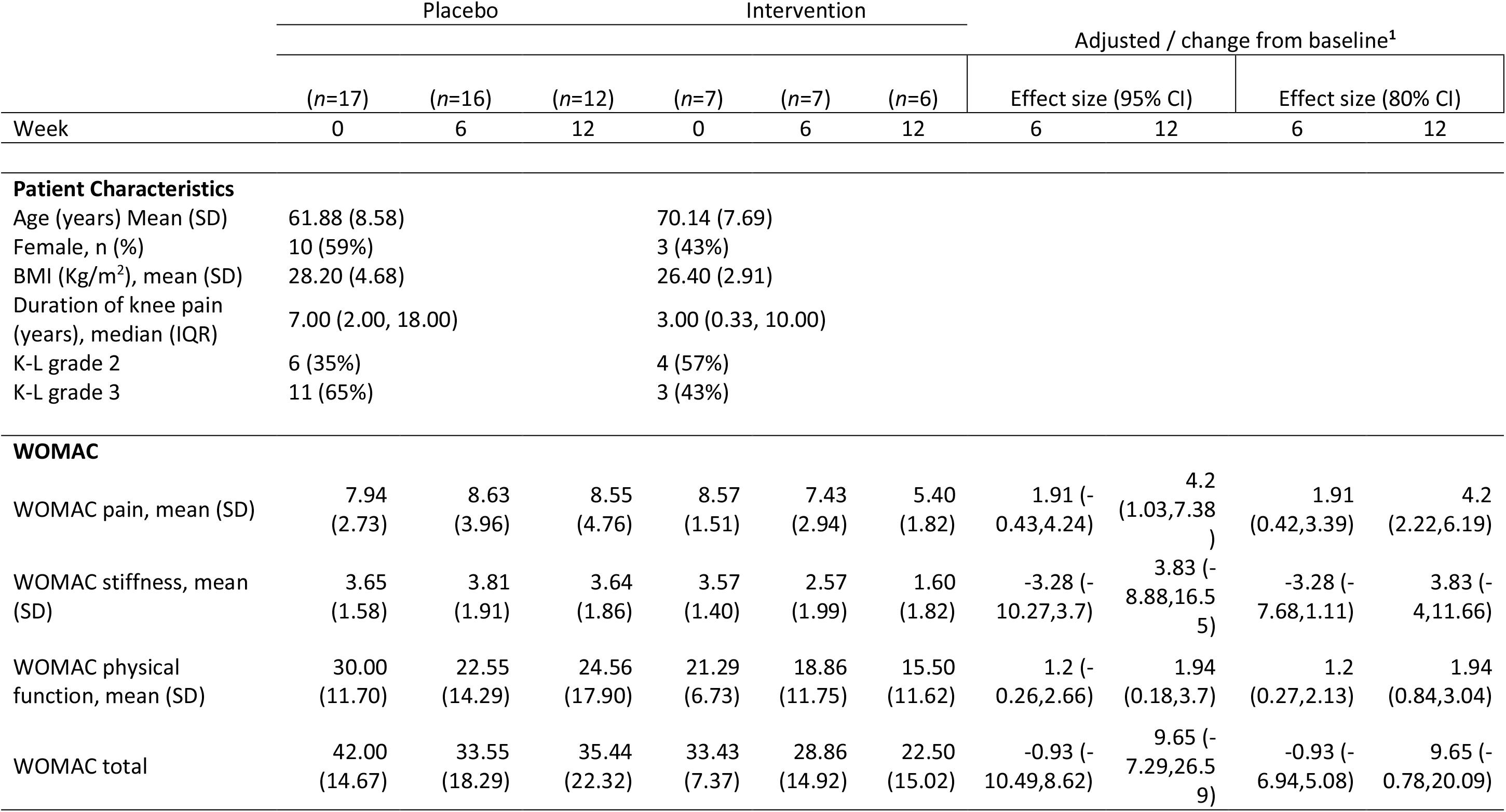
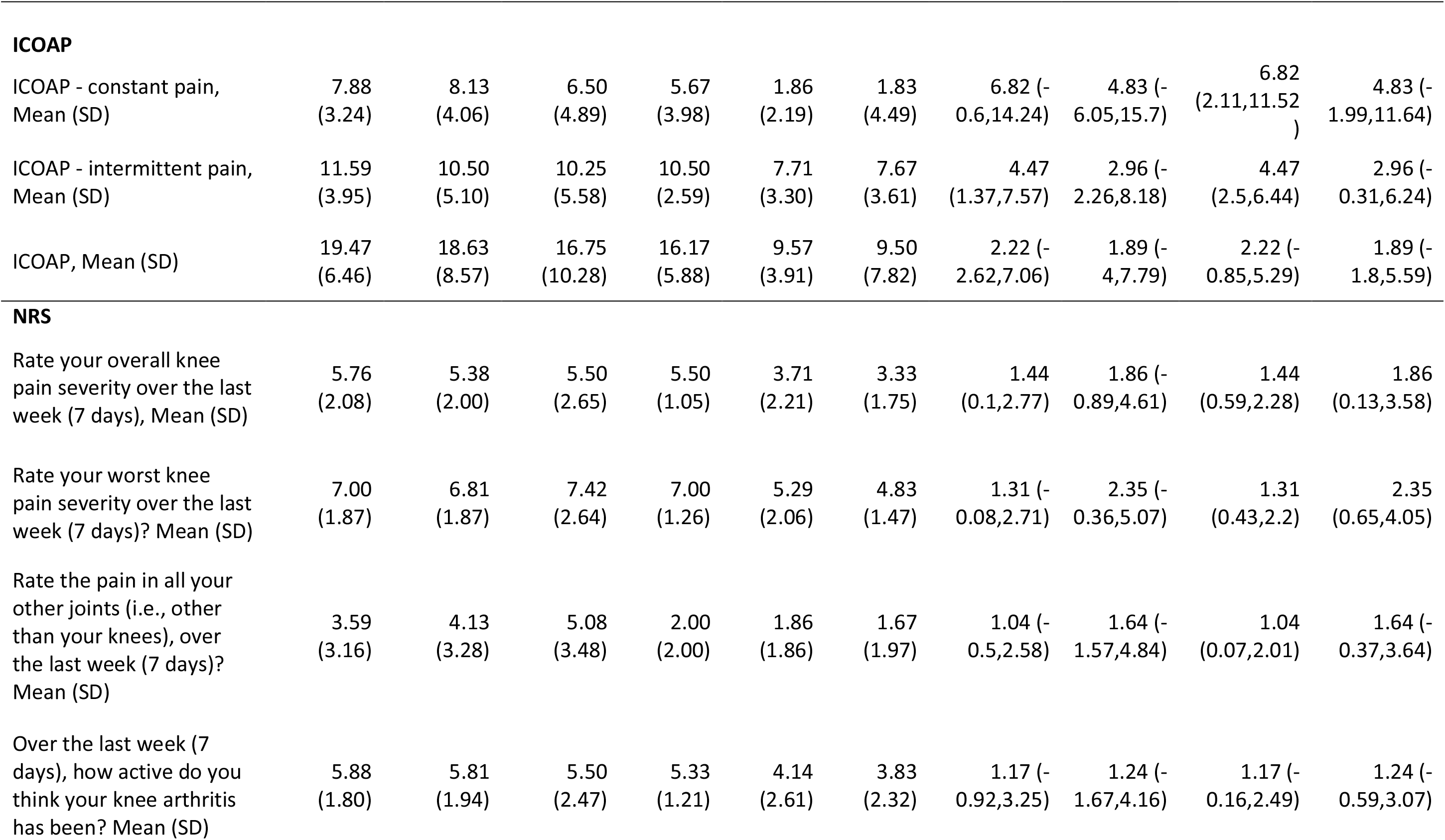
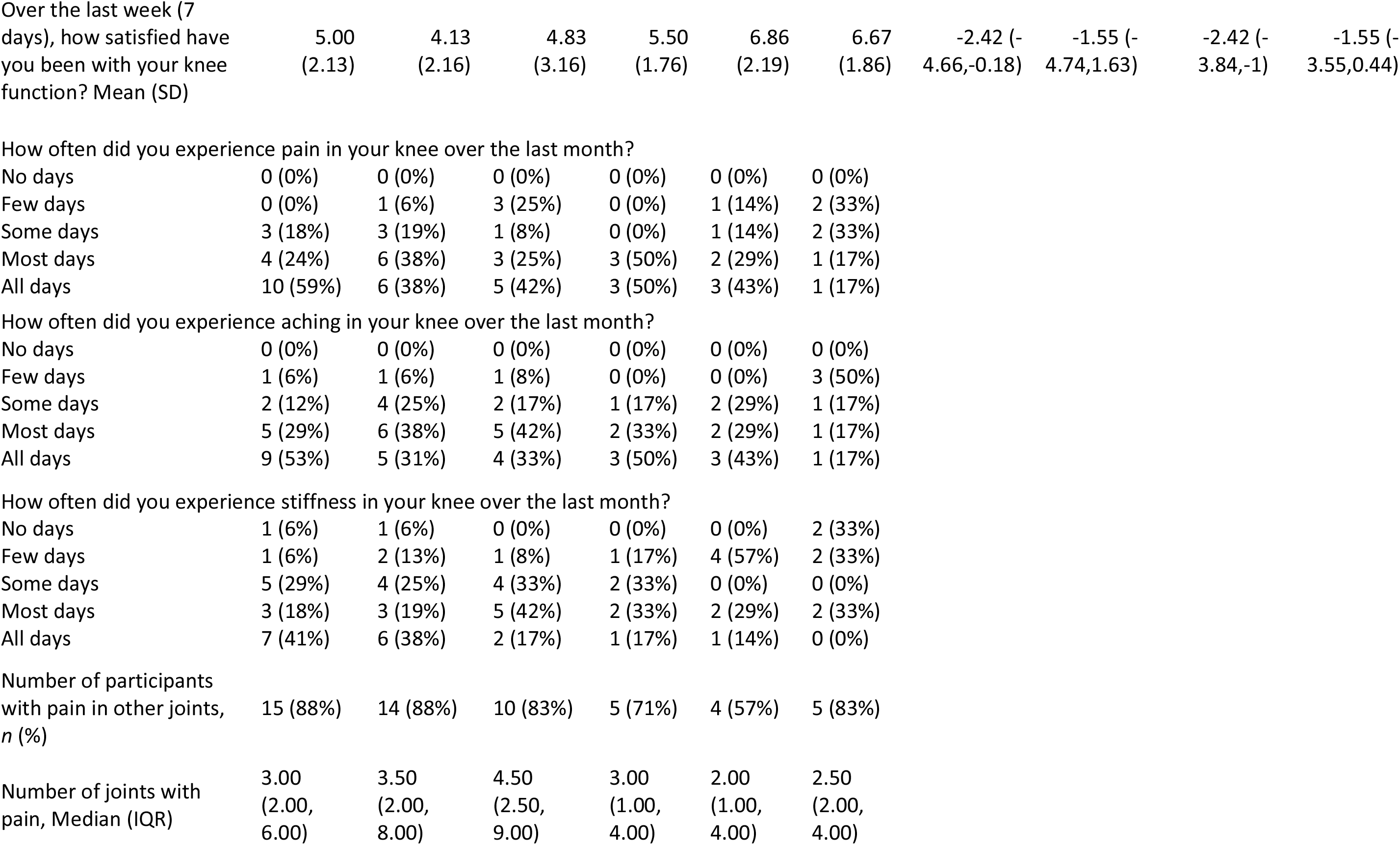
Patient personal and clinical characteristics at baseline (week 0), with reported outcome measures for WOMAC, ICOAP and NRS for 6 and 12 weeks. Data are mean or mean scores (SD), or number of patients (percent) where indicated. Effect size is the adjusted/change from baseline (differences in medians^1^) with 95% confidence intervals.

### Outcome

Primary outcome: The WOMAC pain score was reduced following intervention compared to the placebo group. The effect size (WOMAC pain mean difference from baseline at 12 weeks), for pain scores (scale 0 – 20) was decreased by 4.2 (95% CI: 1.03,7.38). The confidence interval for the difference encompassed the minimal clinically important difference (MCID) (estimated to be 6 units on WOMAC pain scaled 0-100 and equivalent to 1.2 when adjusted here a 0 – 20 scale)^15^ thus providing preliminary evidence of a meaningful treatment effect.

The broccoli intervention showed a clear and common pattern of decreased pain scores over time, across WOMAC, ICOAP and NRS pain measures. ICOAP MCID at weeks 6 and 12 respectively, showed ‘improvement’ for constant pain in the intervention group^16^. A trend for increased satisfaction with knee function was seen in the intervention group. These patterns were not observed in the placebo group. There was no difference in the number of patients with pain in other joints or number of joints with pain (**Table 1**). Self-reported use of rescue analgesics/NSAIDS for knee pain remained constant in both the placebo and intervention arms.

Study compliance and intervention adherence was considered high (>70%-100%) while acceptability for randomisation was 100%. Acceptability of intervention was 92% (22 of 24); two participants did not like the soup.

FFQ dietary data collection was 100%, and 24-hour dietary recall data collection was 100% at all visits except in the control group at week 6 (88%) (data available on request).

Metabolites of glucoraphanin were detected in plasma and urine samples in the intervention group (**Supplementary file 3**).

Participants could not reliably guess their treatment arm (32.7% correct, 23% incorrect, 43% unsure).

### Adverse events

There were 10 adverse events, none were serious, 3 related to the intervention, two of which were related, unexpected and mild, one was related, expected and mild. AE related to the intervention were one episode of vomiting after soup consumption, one report of loose stool and one report of indigestion.

## Discussion

Trial objectives did not include statistical testing for significance in terms of changes for outcome measures given the nature of the feasibility study, however, a clear pattern for a reduction in pain measures across WOMAC, ICOAP and NRS pain subscales was observed in the broccoli intervention group but not the placebo, including an increase in satisfaction for joint function over time. We recruited 37.5% of our target with a 95.7% participant retention and report a high level of compliance (self-reported adherence to protocol and objective measures of metabolites and biomarkers) reflecting high participant motivation and acceptability despite the unprecedented conditions. Dietary data, plasma and 24-hour urine samples were collected where COVID-19 restrictions allowed, enabling the detection of SFN metabolites for objective measures of compliance (**supplementary file 3**). Taken together the results suggest there is significant interest from patients for this type of study, strong commitment from participants and that the study design was feasible under normal circumstances. A food intervention is feasible, and it is important to design trials well. The advent of COVID-19 illuminated the issues of storage space and longevity, but with recent developments in process a way forward has been identified to address this. Food interventions are better to examine than supplements as the population-based data relate to foods rich in glucosinolates, they are a more sustainable approach, environmentally more sensitive, and can be built into the habitual diet paving a way to prevention. SFN is an unstable metabolite of glucoraphanin and difficult to deliver outside of a food matrix since it is well known to require the action of myrosinase present in, for example, broccoli. The next step is to carry out a full-scale trial using a food intervention.

### Limitations of the study

COVID-19 curtailed data collection and restricted sample size below that originally planned, however we remained able to derive meaningful interpretation and meet our original study aims. The study had a short time scale (12 weeks). A longer study would be useful to understand how a long-term intervention might be accepted, important for chronic conditions such as OA. The full sample size fell short of the number anticipated, therefore we were unable to use the data to provide a reliable estimate of sample size for a full trial. A full trial should address impact on long term pain and disease progression since some supplements have been reported to not have clinically important impacts in the medium to longer term^17^. While the protocol asked participants to consume their soup for lunch on the day prior to a visit day, the number of hours from consumption of last soup before sample collection and recording of pain measures was not reliably recorded. This should be standardised in future study. Participants were excluded if they did not like broccoli to maximise compliance and retention, and so a food intervention should account for this in future developments. While most patients tolerated the soups well, two patients withdrew because they did not like the soup.

## Conclusion

We report high participant acceptability, adherence, and retention for the study design. We show that it is feasible to collect dietary and biological samples for objective measures of compliance for the intervention and that blinding was sufficient. Notwithstanding the challenges of underpower we observed clear patterns for improved pain across a range of pain measures and the MCID was met for both the primary outcome and ICOAP constant pain measure. We demonstrate the intervention to be feasible, and a larger trial should be conducted.

## Data Availability

All data produced in the present study are available upon reasonable request to the authors

## Acknowledgements

Our thanks to the study participants for their commitment through a very difficult period, to the study team, research nurses, and doctors; Fiona Brudenell-Straw, Lisa Cook, Lizzy Daniel, Asim Ghouri, Robert Hindmarsh, Kiran Khokar, Teja Kodali, Angela Nauth, Iraklis Papageorgiou, Tracey Swingler, Rabia Thompson, Nicola Ward, and Celia Whitehouse, and Trial committee Simon Donell (Independent Chair), Sam Norton (Independent Biostatistician) and Trish Phillips (Independent member) for their tenacity and team spirit throughout such an unprecedented time.

## Author contributions

Conception and design: R.K.D, A.C^3^, A.C^1^, W.F, I.M, S.R.K, P.G.C, I.M.C, A.M

Analysis and interpretation of the data: R.K.D, G.B, S.S, P.K, A.C^3^, A.C^1^, W.F, I.M, S.R.K, P.G.C, I.M.C, A.M

Drafting of the article: R.K.D, A.C^1^, I.M.C, A.M

Critical revision of the article for important intellectual content: R.K.D, A.C^3^, A.C^1^, W.F, I.M, S.R.K, P.G.C, I.M.C, A.M

Final approval of the article: R.K.D, L.W, G.B, S.S, P.K, A.C^3^, A.C^1^, W.F, I.M, S.R.K, P.G.C, I.M.C, A.M

Provision of study materials or patients: R.K.D, L.W, I.M, S.R.K, P.G.C. Statistical expertise: A.C^1^, A.M

Obtaining of funding: A.C^3^, A.C^1^, W.F, I.M, S.R.K, P.G.C, I.M.C, A.M

Administrative, technical, or logistic support: R.K.D, L.W, S.R.K, Collection and assembly of data: R.K.D, L.W, G.B, S.S, A.C^1^, I.M, S.R.K.

---

**Figure 1.**
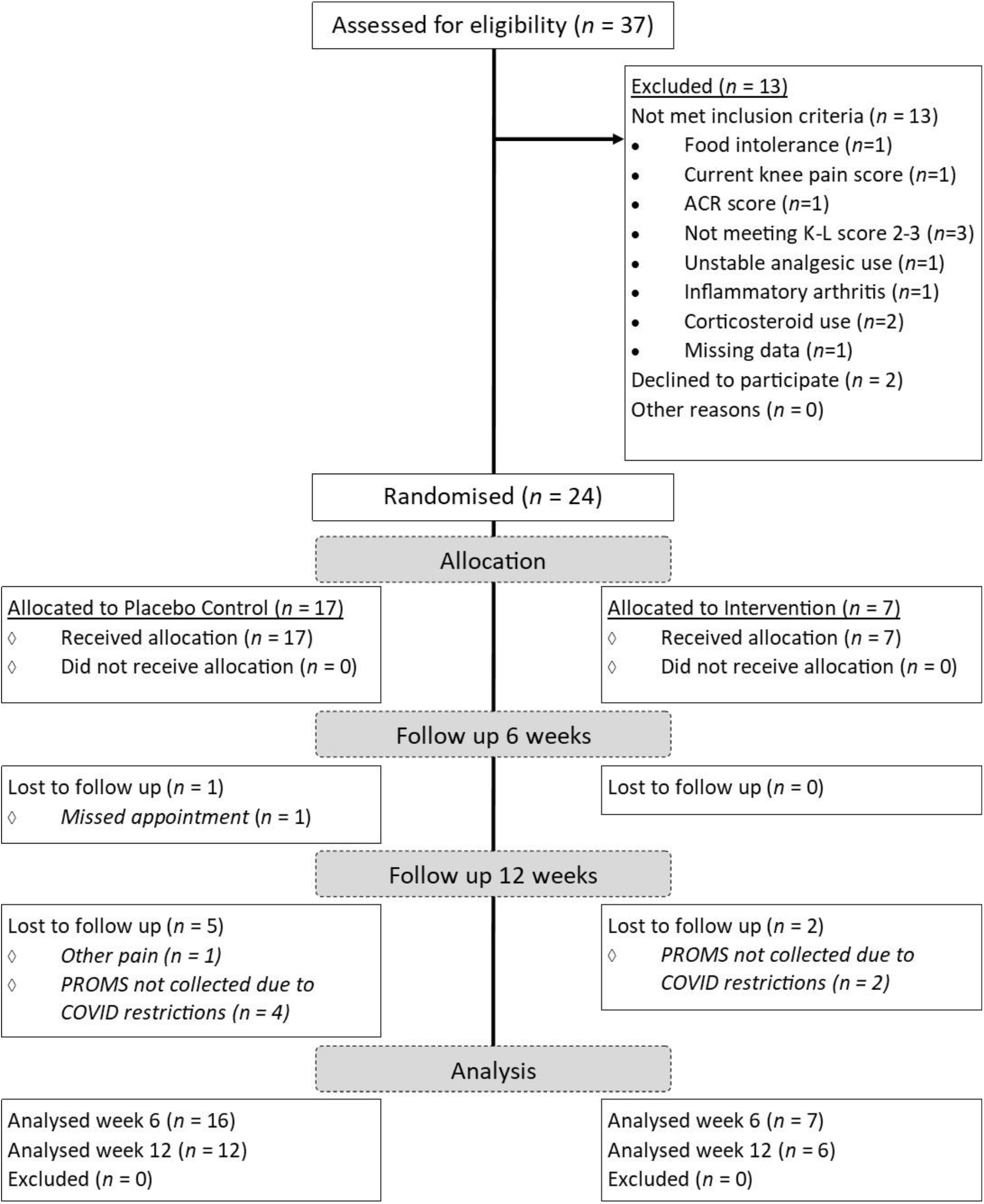
CONSORT flow diagram for BRIO study showing participant flow through screening, randomisation, intervention allocation, follow-up, and analysis.

