## Supplementary file 1 for "The BRoccoli In Osteoarthritis (BRIO study) - A randomised controlled feasibility trial to examine the potential protective effect of broccoli bioactives, (specifically sulforaphane), on osteoarthritis"

### Inclusion and exclusion criteria

**Inclusion Criteria**

• Current knee pain, defined as pain in either knee, in the (one) month before Visit 1, for which the patient gives a severity score of at least 4 on a 0-10 numeric rating scale (NRS).

• The target knee should fulfil the criteria of the American College of Rheumatology (ACR) for knee osteoarthritis defined as knee pain for most days of the prior month and at least one of the following three factors: age over 50 years; morning stiffness of less than 30 minutes; knee crepitus on motion.

• Kellgren Lawrence grade 2-3

• Stable analgesic usage (if using) for 4 weeks prior to trial entry and throughout the trial duration

• Able to adhere to the study visit schedule and other protocol requirements (willing to consume soup intervention).

• Willing to provide 24-hour urine collection samples (x3)

• Capable of giving informed consent and the consent must be obtained prior to any screening procedures.

**Exclusion Criteria**

• The presence of any inflammatory arthritis (e.g., gout, reactive arthritis, rheumatoid arthritis, psoriatic arthritis, seronegative spondyloarthropathy) or fibromyalgia or metabolic bone disease.

• Any clinically significant uncontrolled concurrent illness, which, in the opinion of the Investigator, would impair ability to give informed consent or take part in or complete this clinical study.

• Known or suspected intolerance or hypersensitivity to the investigational product (broccoli) or standardised meal (see section 9.2), closely related compounds, or any of the stated ingredients.

• Use of an investigational product within 30 days prior to 'run in' period or active enrolment in another drug or vaccine clinical study.

• Significant knee injury or any knee surgery within the 6 months preceding enrolment in the study.

• A history of partial or complete joint replacement surgery in the signal knee at any time, listed for knee surgery, or anticipating knee surgery during the study period.

• Poor tolerability of venepuncture or lack of adequate venous access for required blood sampling during the study period

• Nutritional deficiency

• Use of anticoagulant medication (see notes for inclusion exclusion criteria)

• Use of intra-articular hyaluronic acid in the signal knee within the 3 months preceding enrolment in the study.

• Use of intra-articular, intra-muscular or oral corticosteroids in the 2 months preceding enrolment.

• Commencement of non-pharmacological interventions within two months preceding enrolment.

• Persons less than 50 years

• Pregnant/lactating women
