## Supplementary file 2 for "The BRoccoli In Osteoarthritis (BRIO study) - A randomised controlled feasibility trial to examine the potential protective effect of broccoli bioactives, (specifically sulforaphane), on osteoarthritis"

**List of restricted foods**

Participants were asked to avoid the following foods for 3 days prior to the baseline visit and throughout the study period. Participants were given contact details in case they had any questions.

- Broccoli
- Brussel sprouts
- Cress (all varieties)
- Cabbage
- Radish
- Pak choi / bok choy
- Cauliflower
- Horseradish
- Collard greens
- Arugula (rocket)
- Kale
- Swede / rutabaga
- Canola/rapeseed
- Turnip root
- Broccoli sprouts
- Mustard greens
- Kohlrabi
