## Supplementary file 3 for "The BRoccoli In Osteoarthritis (BRIO study) - A randomised controlled feasibility trial to examine the potential protective effect of broccoli bioactives, (specifically sulforaphane), on osteoarthritis"

### BRIO Study Blood and Urine Metabolites

| | Baseline | | 6 weeks | | | | | | 12 weeks | | | | | |
|---|---|---|---|---|---|---|---|---|---|---|---|---|---|---|
| Outcome | Placebo (n=17) | Intervention (n=7) | Placebo (n=16) | Intervention (n=7) | Unadjusted | | Adjusted / change from baseline1 | | Placebo (n=12) | Intervention (n=6) | Unadjusted | | Adjusted / change from baseline1 | |
| Sample | (n=17) | (n=7) | N=12 | N=5 | Effect size (95% CI) | Effect size (80% CI) | Effect size (95% CI) | Effect size (80% CI) | N=8 | N = 3 | Effect size (95% CI) | Effect size (80% CI) | Effect size (95% CI) | Effect size (80% CI) |
| SF plasma (nM), Median (IQR) | 0.71 (0.59, 0.98) | 0.55 (0.53, 0.60) | 0.49 (0.48, 54.75) | 7.90 (1.00, 227.26) | 7.36 (-107.94,1777.76) | 7.36 (-0.00,226.78) | -7.37 (-1779.86,105.85) | -7.37 (-226.97,-0.15) | 0.51 (0.48, 0.53) | 0.80 (0.51, 10.29) | 0.29 (-0.03,9.79) | 0.29 (-0.01,9.77) | -0.82 (-9.98,0.00) | -0.82 (-9.90,-0.15) |
| SF urine (nM x10$^{-1}$), Median (IQR) | 0.46 (0.43, 0.48) | 0.46 (0.44, 0.55) | 0.33 (0.28, 0.36) | 0.40 (0.37, 0.47) | 0.09 (0.02,1.57) | 0.09 (0.04,0.15) | -0.02 (-1.59,0.11) | -0.02 (-0.24,0.07) | 0.29 (0.28, 0.30) | 0.39 (0.33, 1.82) | 0.10 (0.03,1.54) | 0.10 (0.04,1.53) | -0.10 (-1.55,0.12) | -0.10 (-1.51,0.06) |
| E-NAC (nM), Median (IQR) | 0.11 (0.07, 0.41) | 0.23 (0.10, 0.98) | 0.11 (0.05, 0.13) | 2.35 (0.45, 4.54) | 2.24 (0.06,7.90) | 2.24 (0.31,4.49) | -1.24 (-7.88,0.01) | -1.24 (-3.68,-0.13) | 0.11 (0.10, 0.14) | 1.00 (0.07, 4.76) | 0.89 (-0.08,4.66) | 0.89 (-0.03,4.63) | -0.10 (-4.80,0.29) | -0.10 (-4.67,0.24) |
| SF-Cys (nM), Median (IQR) | 0.38 (0.17, 0.70) | 0.50 (0.25, 0.58) | 0.54 (0.41, 0.76) | 2.14 (1.36, 3.51) | 1.60 (-0.07,3.60) | 1.60 (0.41,3.07) | -1.51 (-3.75,-0.29) | -1.51 (-2.81,-0.58) | 0.51 (0.42, 0.81) | 1.18 (0.41, 2.38) | 0.67 (-0.35,1.90) | 0.67 (-0.07,1.84) | -0.40 (-2.64,0.26) | -0.40 (-2.39,0.21) |
| SF-NAC (nM x10$^{-1}$), Median (IQR) | 0.93 (0.80, 1.39) | 1.02 (0.86, 1.57) | 0.82 (0.80, 0.89) | 6.03 (1.17, 6.17) | 5.19 (-0.01,41.43) | 5.19 (0.32,5.36) | -3.38 (-41.54,0.13) | -3.38 (-4.98,-0.01) | 0.81 (0.78, 0.87) | 4.17 (2.43, 25.17) | 3.36 (1.58,24.38) | 3.36 (1.65,24.34) | -2.96 (-24.90,-1.19) | -2.96 (-24.58,-1.49) |

[1] Differences in medians

Samples missing due to COVID restrictions
